# High Prevalence of Resistance to First-Line Drugs and Multidrug Resistance in Patients with Tuberculosis and Type 2 Diabetes Mellitus at a Referral Hospital in Peru

**DOI:** 10.1101/2025.11.06.25339704

**Authors:** Sergio Barboza-Panaifo, Italo Erazo Cárdenas, Nallely Chapoñan-Agip, Anabell Tenorio-Quispe, Marlon Yovera-Aldana

**Author notes:** Sergio Barboza-Panaifo, Italo Erazo-Cárdenas, Nallely V. Chapoñan-Agip, Anabell M Tenorio-Quispe, Marlon Yovera-Aldana. Corresponding autor: Marlon Yovera-Aldana.

## Abstract

**Introduction:** The coexistence of tuberculosis (TB) and type 2 diabetes mellitus (T2DM) poses an emerging public health challenge, especially in high-TB-burden settings such as Peru. This comorbidity has been linked to unfavorable clinical outcomes and a higher likelihood of developing drug-resistant TB.

**Objective:** To estimate the prevalence of resistance to first-line antituberculosis drugs and identify factors associated with multidrug resistance (MDR) among patients with T2DM and pulmonary TB treated at a public hospital in Lima, Peru.

**Methods:** A cross-sectional study was conducted using secondary data from 130 adults with T2DM and microbiologically confirmed pulmonary TB managed at Hospital María Auxiliadora between 2015 and 2019. Drug susceptibility results were categorized as sensitive, monoresistant, polyresistant, or MDR. Factors associated with MDR and resistance to at least one drug were evaluated using Poisson regression models with robust variance.

**Results:** The prevalence of resistance to at least one first-line drug was 34.6%, and the prevalence of MDR-TB was 21.5%. The most frequent resistances were to isoniazid (31%) and rifampicin (27%). In the multivariable analysis, age ≥70 years (PR = 4.13; 95% CI: 1.16–14.7) and age 40–49 years (PR = 2.99; 95% CI: 1.00–8.97) were independently associated with MDR-TB. Furthermore, a T2DM duration ≥5 years was significantly associated with resistance to at least one drug (adjusted PR ≈ 2.1).

**Conclusion:** Patients with T2DM and pulmonary TB show a high prevalence of drug resistance, including MDR-TB. Older age and longer duration of diabetes are significant risk factors. These findings highlight the need for integrated TB–diabetes management strategies to strengthen early detection and control of drug-resistant TB in this vulnerable population.

## Introduction

On a global scale, tuberculosis (TB) has shown a sustained decline in both incidence and overall mortality since 1990, largely attributable to the implementation of the Directly Observed Treatment, Short-Course (DOTS) strategy. Nevertheless, this decrease has been less pronounced in developing regions (1,2). In Latin America, TB incidence has also declined, although significant challenges persist. In Brazil, the incidence decreased from 42.8 to 35.2 cases per 100,000 population between 2001 and 2017, but rose again to 39.8 cases in 2023, particularly affecting vulnerable populations, a figure that exceeds the World Health Organization (WHO) target of 6.7 per 100,000 population (6,7). Peru accounts for 14% of all TB cases in South America, with Lima Metropolitan Area and Callao reporting 55.6% of national cases (9). Despite reductions in new cases in previous decades (8), the persistently high burden in specific regions underscores the need for tailored public health strategies.

One contributing factor to this limited progress is the rising resistance to first-line anti-tuberculosis drugs (11,12). Resistance patterns vary acoording to the number of affected drugs and their therapeutic relevance. Multidrug-resistant tuberculosis (MDR-TB), defined as resistance to both isoniazid and rifampicin, is particularly associated with poorer clinical outcomes. In a region of central Peru, the prevalence of MDR-TB was estimated at 3.7%, with 46% of these cases having received prior treatment and 59% experiencing treatment failure (13). In the capital city of Peru, another study reported an MDR-TB prevalence of 13% (14), reflecting a substantial disease burden driven by persistent, inadequately managed cases.

The coexistence of TB with chronic diseases such as diabetes mellitus (DM) is substantial. Immunometabolic alterations induced by sustained hyperglycemia increase susceptibility to *Mycobacterium tuberculosis* infection and lead to suboptimal therapeutic responses, potentially fostering the emergence of drug-resistant strains (15,16) Globally, the estimated incidence and prevalence of TB among individuals with DM are 129 and 511 cases per 100,000 person-years, respectively (18). In the capital city of Peru, 12% of MDR-TB cases occur among patients with DM, and this proportion rises to 28% among those with a history of previous TB treatment (19). Given that prior TB episodes are common among patients with DM and contribute to the development of resistance, this study aimed to characterize bacterial resistance patterns and identify associated factors, distinguishing between any drug resistance and multidrug resistance, among adults with DM treated in a public hospital in Peru.

## Material and Methods

### Study design and setting

We conducted a cross sectional, descriptive study based on a secondary analysis of a clinical database from the” Divino Niño” Center of Excellence for Tuberculosis at Maria Auxiliadora Hospital (HMA) in Lima, Peru. This high-capacity referral center is part of the Peruvian Ministry of Health and serves patients from 118 primary care facilities across southern Lima. The population covered is predominantly of low socioeconomic status and receives subsidized public health insurance. The Center of Excellence manages complex tuberculosis cases, including drug resistant TB and TB with comorbidities. Data collection and analysis were carried out during the second half of 2021.

### Study population, sample, and sampling strategy

Patients aged 18 years or older with a diagnosis of type 2 diabetes mellitus (T2DM) were included. The diagnosis was confirmed based on medical history, self-report, use of hypoglycemic agents, or diagnosis at admission according to the American Diabetes Association (ADA) criteria. In addition, participants were required to have active pulmonary tuberculosis confirmed by drug susceptibility testing for anti-tuberculosis agents, performed within the first two months of treatment. Furthermore, only patients treated between 2015 and 2019 were considered. Patients with extrapulmonary tuberculosis, HIV infection, or inconsistent clinical records were excluded. We conducted a census of all eligible patients treated during the study period; no sampling was performed.

### Variables

#### Resistance classification

Bacterial resistance was defined according to the number and therapeutic hierarchy of the four main anti-tuberculosis drugs, and was classified into four categories. Multidrug-resistant tuberculosis (MDR-TB) was defined as resistance to both isoniazid and rifampicin. Oligoresistance was defined as resistance to more than two drugs, excluding the combination of isoniazid and rifampicin. Monoresistance was defined as resistance to any one of the four drugs in isolation, and drug-susceptible tuberculosis was defined as susceptibility to all four drugs. Data were extracted by endocrinology service physicians from reports provided by the National Institute of Health (INS) and documented in the patients’ medical records. The INS performs drug susceptibility testing (DST) in accordance with World Health Organization (WHO) criteria and the 2019 Peruvian Technical Standard. For the isolation of *Mycobacterium tuberculosis*, the absolute concentration method on Löwenstein–Jensen medium is used. The concentrations used for susceptibility testing were as follows: isoniazid, 0.2 μg/mL; rifampicin, 40 μg/mL; streptomycin, 10 μg/mL; and ethambutol, 2 μg/mL. For the purposes of this study, two primary outcomes were defined. The first was resistance to any first-line drug (monoresistance, multidrug resistance, or oligoresistance classified as “yes,” and drug-susceptible cases classified as “no”). The second was multidrug resistance (yes or no).

#### Other variables

Sociodemographic variables were categorized as follows: age (23–39 years, 40–49 years, 50–59 years, 60–69 years, and 70–85 years); sex (male, female); educational level (primary/secondary, higher); and place of residence (San Juan de Miraflores, Villa María del Triunfo, Villa El Salvador, Chorrillos, and other districts). Clinical variables included: duration of diabetes diagnosis (≤5 years, ≥5 years); body mass index (BMI) (<25 kg/m², 25–29.9 kg/m², ≥30 kg/m²); history of tuberculosis (yes, no); diabetes treatment regimen (diet only, metformin only, basal insulin ± metformin, insulin only); and adherence to hypoglycemic treatment, defined as compliance with the prescribed dosage (yes, no). Fasting plasma glucose levels (<130 mg/dL, 130–200 mg/dL, ≥200 mg/dL), postprandial glucose levels (<140 mg/dL, 140–180 mg/dL, ≥180 mg/dL), and glycemic control according to HbA1c (≤7%, 7–9%, ≥9%) from the most recent month on record were also described.

### Procedures and Analysis Plan

Approval was obtained from the Office for Research and Teaching Support of the HMA, as well as from the coordination of the Endocrinology Service. Access to the database was granted on August 1, 2021, and the analysis was conducted through December 31, 2021.

Data were exported from Excel to STATA statistical software, version 16.0 (StataCorp, College Station, Texas, USA).

For the analysis, categorical variables (sex, educational level, BMI, diabetes treatment regimen, treatment adherence, glycated hemoglobin, resistance to tuberculosis treatment, history of pulmonary tuberculosis, and resistance to anti-tuberculosis treatment) were summarized using absolute and relative frequencies. Age and HbA1c were summarized using the mean and standard deviation, whereas duration of disease was presented as the median and interquartile range due to its non-normal distribution.

Associations between demographic and clinical variables and the two primary outcomes, resistance to any first-line drug and multidrug resistance, were assessed. Pearson’s chi-square test was used for categorical variables. When more than 20% of the expected values were less than 5, Fisher’s exact test was applied.

In the multivariable analysis, a generalized linear model with a log link and robust Poisson variance was applied—one model for resistance to any first-line drug and another for multidrug resistance. This approach was chosen due to the high prevalence of the outcome (>10%), which could overestimate the odds ratio in traditional logistic regression models. Crude and adjusted prevalence ratios (PRs) with their 95% confidence intervals were obtained. Variable selection for adjustment was based on a statistical model, including those with a *p*-value <0.2 due to the exploratory nature of the study. The models included a maximum of three to four variables, given the number of subjects, to avoid overfitting. Multicollinearity analysis among covariates was performed in the adjusted models.

Sample adequacy was evaluated to estimate the prevalence of multidrug-resistant tuberculosis (MDR-TB) and any first-line drug resistance among patients with T2DM and pulmonary TB. Using EPIDAT version 4.2, the estimated precision for the prevalence of MDR-TB (21.5%) was ±7.1%, and for resistance to at least one first-line drug (34.6%) was ±8.2%, assuming a 95% confidence level in a sample of 130 subjects (20).

### Ethics statement

The research protocol was approved by the Ethics Committees of María Auxiliadora Hospital (HMA/CIEI/0016/2021) and Universidad Científica del Sur (065-2021-PRE15). The data used in this study were obtained from the Endocrinology Department and did not contain any personally indentifiable information.

## Results

### Subject Selection

From 2015 and 2019, a total of 360 patients diagnosed with both diabetes and tuberculosis were treated. After applying the eligibility criteria in the database, a total of 130 subjects were included (**Fig 1**).

**Fig 1.**
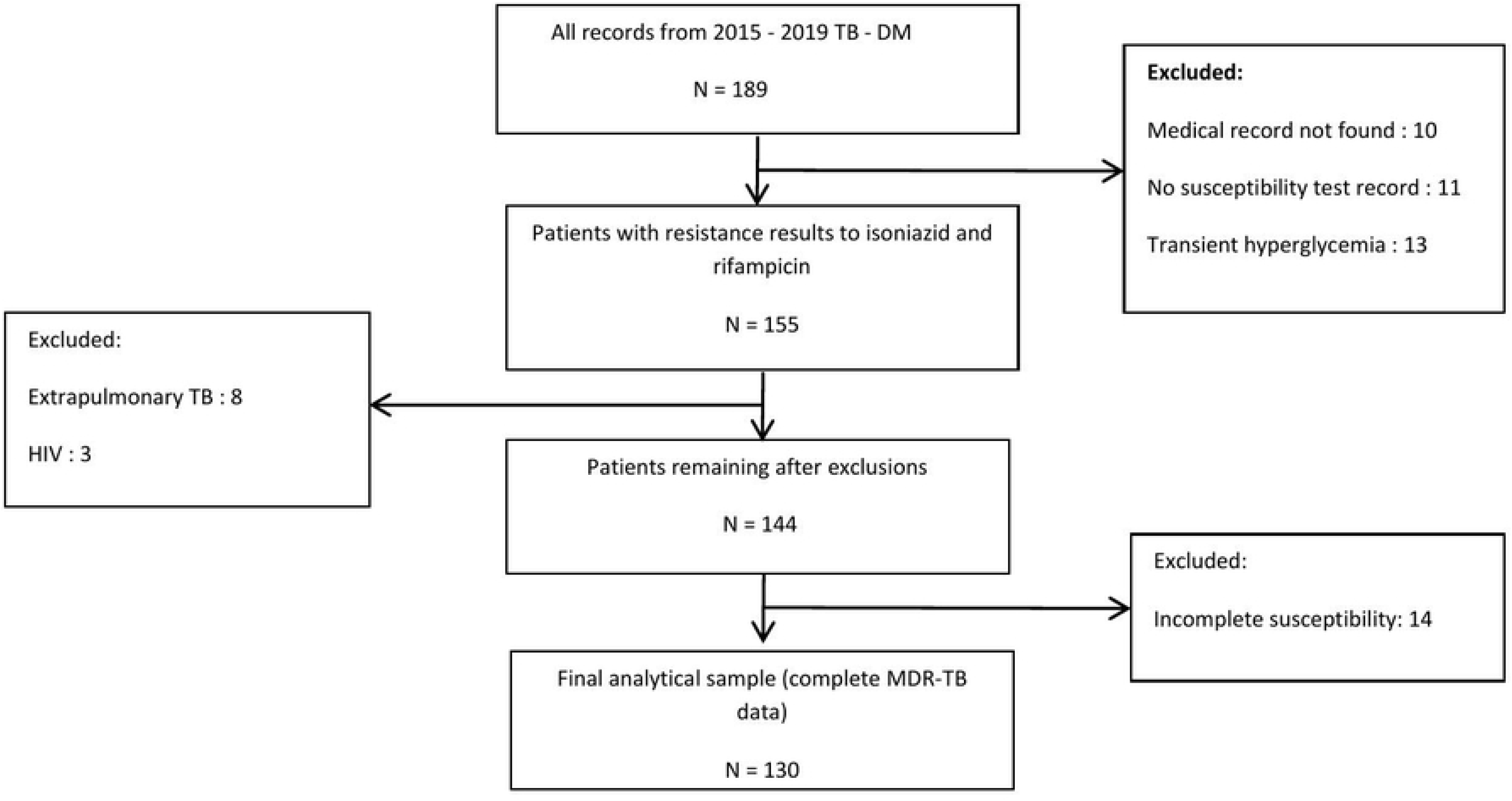
**Flow diagram of patient selection for the study on multidrug-resistant tuberculosis (MDR-TB) and type 2 diabetes mellitus.**

Among the 130 patients with complete data, 28 (21.5%) had MDR-TB and 45 (34.6%) showed resistance to at least one first-line drug

### General Characteristics

Of the sample included, 54.6% were male, and the largest age group was between 50 and 59 years old, accounting for 33.1%. The majority of participants (54.6%) attended primary or secondary school.

A clinical evaluation revealed that 36.7% of individuals were overweight and 13.9% were obese. Additionally, 79.2% of participants had a history of tuberculosis, and 41.7% had diabetes for < 5 years. As diabetes treatment, 55.1% of participants used the basal insulin regimen and 35.4% used the basal-bolus regimen.

Regarding glycemic control, 86.6% had fasting blood glucose levels ≥130 mg/dL and 88.9% had postprandial blood glucose levels ≥180 mg/dL. For glycosylated hemoglobin, 15.8% had values between 7% and 9%, while 71.1% had a value ≥ 9% **(Table 1).**

**Table 1.** Sociodemographic and Clinical Characteristics of Patients with Type 2 Diabetes Mellitus and Pulmonary Tuberculosis.

| Characteristics | Total sample<br>(N=144) | Patients with complete<br>data for MDR<br>(N=130) |
| --- | --- | --- |
| **Demographics** |  |  |
| **Sex** |  |  |
| Male | 78 / 144 (54.1) | 71/130 (54.6) |
| Female | 66 / 144 (45.9) | 59/130 (45.4) |
| **Age, years** |  |  |
| 23- 39 | 25 /144 (17.1) | 25 /130 (19.2 ) |
| 40- 49 | 34 / 144 (23.3) | 31/130 (23.9 ) |
| 50- 59 | 52 / 144 (35.6) | 43/130 (33.1 ) |
| 60- 69 | 26 / 144 (17.8) | 23 /130 (17.7 ) |
| 70 - 85 | 9 / 144 (6.2) | 8 /130 (6.2 ) |
| Mean ± SD | 51,2 ± 11,8 | |
| **Education level** |  |  |
| Primary - secondary | 86 / 144 (59.6) | 71/130 (54.6 ) |
| Higher education | 58 / 144 (40.4) | 59/130 (45.4 ) |
| **Residence** |  |  |
| San Juan de Miraflores | 43 / 144 (30.1) | 40 /130 (30.8) |
| Villa María del Triunfo | 38 / 144 (26.7) | 31 /130 (23.9) |
| Villa el Salvador | 27 / 144 (18.4) | 25/130 (19.2) |
| Chorrillos | 17 / 144 (11.6) | 16/130 (12.3) |
| Other | 19 / 144 (13.2) | 18/130 (13.9) |
| **Clinical** |  |  |
| **Body Mass Index** |  |  |
| Underweight (< 18.5 kg/m<sup>2</sup>) | 19 / 144 (13.1) | 17/130 (13.1 ) |
| Normal (18.5–24.9 kg/m<sup>2</sup>) | 58 / 144 (40.0) | 48/130 (36.9 ) |
| Overweight (25.0–29.9 kg/m<sup>2</sup>) | 49 / 144 (33.8) | 47/130 (36.2 ) |
| Obese (≥ 30 kg/m<sup>2</sup>) | 19 / 144 (13.1) | 18/130 (13.9 ) |
| **Prior tuberculosis** |  |  |
| Present | 118 / 144 (80.8) | 27 /130 (79.2) |
| Absent | 28 / 144 (19.2) | 103 /130 (20.8) |
| **Duration of diabetes, years** |  |  |
| <5 years | 62 / 144 (43.4) | 53 /127 (41.7 ) |
| 5-9.9 years | 44 / 144 (30.8) | 39 /127 (30.7 ) |
| ≥ 10 years | 37 / 144 (25.9) | 35 /127 (27.6 ) |
| Median [IQR] | 5 [2 a 10] |  |
| **Diabetes treatment regimen** |  |  |
| Diet only | 7 / 144 (4.9) | 6 /127 (4.7) |
| Metformin only | 6 / 144(4.2) | 6/127 (4.7) |
| Basal insulin ± metformin | 74 / 144 (51.8) | 70/127 (55.1) |
| Basal insulin -bolus | 56 / 144 (39.1) | 45/127 (35.4) |
| **Adherence to hypoglycemic treatment** |  |  |
| Adherent | 96 / 120 (80.0) | 95 /118 (80.5) |
| Non-adherent | 24 / 120 (20.0) | 23/118 (19.5) |
| **Fasting blood glucose** |  |  |
| <130 mg/dL | 18 / 141 (12.7) | 17/127 (13.4) |
| 130-200 mg/dL | 47 / 141 (33.3) | 40/127 (31.5) |
| ≥ 200 mg/dL | 76 / 141 (54.0) | 70/127 (55.1) |
| **Postprandial blood glucose** |  |  |
| <140 mg/dL | 6 / 120 (5.0) | 6/117 (5.1) |
| 140-180 mg/dL | 7 / 120 (6.0) | 7 /117 (6.0) |
| ≥180 mg/dL | 107 / 120 (89.0) | 104/117 (88.9) |
| **HbA1c** |  |  |
| <7% | 17 / 128(13.3) | 15/114 (13.2) |
| 7-9% | 20 / 128 (15.6) | 18/114 (15.8) |
| ≥9% | 91 / 128 (71.1) | 81/114 (71.1) |
| Mean ± SD | 10.7 ± 3.0 |  |

Data are presented as n / N (%), unless otherwise indicated. N represents the number of patients with available data for each variable. The total number of included patients was 144, of which 130 had complete data for multidrug resistance (MDR) analysis “Primary or secondary” includes participants with no formal education, preschool, primary, or secondary schooling. “Other districts” include Surco, Barranco, Lurín, Pachacámac, San Bartolo, Pucusana, Punta Negra, Punta Hermosa, and Santa María del Mar.

### Prevalence of bacterial resistance to first-line drugs

The drug with the highest resistance was isoniazid with 28.4% (95% CI=21.9–37.0), followed by rifampicin with 26.2% (95% CI=18.8–34.6). Likewise, the prevalence of resistance to some first-line drug was 34.6% (95% CI=26.5–43.4). The respective percentages of multidrug, polyresistance, and monoresistance were 21.5% (95% CI= 14.8–29.6), 2.3% (95% CI= 0.4–6.5), and 10.8% (95% CI= 6.0–17.4) (Table 2).

**Table 2.** Resistance to First-Line Antituberculosis Drugs in Patients with Type 2 Diabetes Mellitus.

|  | n | Prevalence (95% IC) |
| --- | --- | --- |
| **Single drug resistance** |  |  |
| Isoniazid (H ) | 37 | 28.4 (21.9 – 37.0) |
| Rifampicin (R) | 34 | 26.2 (18.8 – 34.6) |
| Pyrazinamide (Z) | 17 | 13.1 (7.8 – 20.1) |
| Ethambutol (E) | 17 | 13.1 (7.8 – 20.1) |
| **Any resistance to ≥1 drug** | 45 | 34.6 (26.5 – 43.4) |
| **Multidrug-resistant TB (MDR-TB)** | 28 | 21.5 (14.8 – 29.6) |
| **(resistance to at least H + R)** |  |  |
| H+R | 13 |  |
| H+R+E | 2 |  |
| H+R+Z | 2 |  |
| H+R+E+Z | 11 |  |
| **Polyresistance** | 3 | 2.3 (0.4 – 6.5) |
| **(resistance to >2 drugs, excluding H + R)** |  |  |
| H+E+Z | 3 |  |
| Other combinations (H + E, H + Z, R + E, R + Z, E + Z, R + E + Z) | 0 |  |
| **Monoresistance** | 14 | 10.8 (6.0 – 17.4) |
| H only | 6 |  |
| R only | 6 |  |
| E only | 1 |  |
| Z only | 1 |  |

Data are expressed as number of cases (n) and prevalence (%; 95% confidence interval). MDR-TB was defined as concurrent resistance to isoniazid (H) and rifampicin (R), with or without resistance to other first-line drugs (E, Z). Polyresistance was defined as resistance to more than two first-line drugs, excluding the H + R combination. Monoresistance corresponds to resistance to a single first-line drug.

### Factors Associated with TB-MDR

In the bivariate analysis, the 50–59 age group had the lowest prevalence of MDR (9.3%). A J-shaped curve was observed, with high prevalence at the extremes, particularly among individuals over 70 years of age (p = 0.039). No differences were observed between categories in the other variables (Table 3).

**Table 3.** Sociodemographic and Clinical Characteristics Associated with Multidrug-Resistant.

| Characteristics | MDR-TB<br>positive<br>n (%) | MDR-TB<br>negative<br>n (%) | p-value | Any drug<br>resistance<br>n (%) | Fully<br>Susceptible<br>n (%) | p-value |
| --- | --- | --- | --- | --- | --- | --- |
| Sex , (n=130) |  |  |  |  |  |  |
| Male | 15 (21.1) | 56(78.9) | 0.900<sup>a</sup> | 22 (31.0) | 49 (69.0) | 0.340<sup>a</sup> |
| Female | 13 (22.0) | 46(78.0) |  | 23 (38.9) | 36 (61.0) |  |
| Age group, years (n=130) |  |  |  |  |  |  |
| 23 - 39 | 7 (28.0) | 18 (72.0) | 0.039<sup>a</sup> | 10 (40.0) | 15 (60.0) | 0.219<sup>a</sup> |
| 40 - 49 | 9 (29.0) | 22 (71.0) |  | 12 (38.7) | 19 (61.3) |  |
| 50 - 59 | 4 (9.3) | 39 (90.7) |  | 9 (20.9) | 34 (79.1) |  |
| 60 - 69 | 4 (17.4) | 19 (82.6) |  | 10 (43.5) | 13 (56.6) |  |
| 70 - 85 | 4 (50.0) | 4 (50.0) |  | 4 (50.0) | 4 (50.0) |  |
| Education level, (n=130) |  |  |  |  |  |  |
| Primary or secondary | 13 (18.3) | 58 (81.7) | 0.326<sup>a</sup> | 20 (28.2) | 51 (71.8) | 0.090<sup>a</sup> |
| Higher education | 15 (25.4) | 44 (74.6) |  | 25 (42.3) | 34 (57.6) |  |
| Distric of residence (n=130) |  |  |  |  |  |  |
| San Juan de Miraflores | 9 (22.5) | 31 (77.5) | 0.926 <sup>a</sup> | 13 (32.5) | 27 (67.5) | 0.845 <sup>a</sup> |
| Villa María del Triunfo | 8 (25.8) | 23 (74.2) |  | 12 (38.7) | 19 (61.3) |  |
| Villa el Salvador | 4 (16.0) | 21 (84.0) |  | 7 (28.0) | 18 (72.0) |  |
| Chorrillos | 3 (18.8) | 13 (81.3) |  | 7 (43.8) | 9 (56.3) |  |
| Other* | 4 (22.2) | 14 (77.8) |  | 6 (33.3) | 12 (66.7) |  |
| **Body mass index, (n=130)** |  |  |  |  |  |  |
| Underweight (< 18.5 kg/m<sup>2</sup>) | 7 (41.2) | 10 (58.8) | 0.120 <sup>a</sup> | 7 (41.2) | 10 (58.8) | 0.550 <sup>a</sup> |
| Normal (18.5–24.9 kg/m<sup>2</sup>) | 7 (14.6) | 41 (85.4) |  | 17 (35.4) | 31 (64.6) |  |
| Overweight (25.0–29.9 kg/m<sup>2</sup>) | 9 (19.2) | 38 (80.8) |  | 13 (27.7) | 34 (72.3) |  |
| Obese (≥ 30 kg/m<sup>2</sup>) | 5 (27.8) | 13 (72.2) |  | 8 (44.4) | 10 (55.6) |  |
| **Duration of diabetes (years), (n=127)** |  |  |  |  |  |  |
| <5 | 7 (13.2) | 46 (86.8) | 0.193 <sup>a</sup> | 11 (20.8) | 42 (79.3) | 0.031 <sup>a</sup> |
| 5 a 9.9 | 9 (23.1) | 30 (76.9) |  | 17 (43.6) | 22 (56.4) |  |
| ≥ 10 | 10 (28.6) | 25 (71.4) |  | 15 (42.9) | 20 (57.1) |  |
| **History of Tuberculosis (n=130)** |  |  |  |  |  |  |
| Present | 22 (21.3) | 81 (78.6) | 0.923 <sup>a</sup> | 9 (33.3) | 18 (66.7) | 0.875 <sup>a</sup> |
| Absent | 6 (22.2) | 21 (77.8) |  | 36 (34.9) | 67 (65.1) |  |
| **Type of diabetes treatment (n=127)** |  |  |  |  |  |  |
| Diet only | 1 (16.7) | 5 (83.3) | 0.907 <sup>b</sup> | 2 (33.3) | 4 (66.7) | 0.797 <sup>b</sup> |
| Metformin only | 1 (16.7) | 5 (83.3) |  | 1 (16.7) | 5 (83.3) |  |
| Basal insulin | 16 (22.9) | 54 (77.1) |  | 26 (37.1) | 44 (62.9) |  |
| Basal + prandial insulin | 8 (17.8) | 37 (82.2) |  | 14 (31.1) | 31 (68.9) |  |
| **Adherence to hypoglycemic treatment (n=118)** |  |  |  |  |  |  |
| Adherent | 16 (16.8) | 79 (83.2) | 0.140 <sup>a</sup> | 10 (43.5) | 13 (56.5) | 0.236 <sup>a</sup> |
| Non-adherent | 7 (30.4) | 16 (69.6) |  | 29 (30.5) | 66 (69.5) |  |
| **Fasting blood glucose, (n=117)** |  |  |  |  |  |  |
| < 130mg/dL | 1 (5.9) | 16 (94.1) | 0.277 <sup>a</sup> | 6 (40.0) | 9 (60.0) | 0.407 <sup>a</sup> |
| 130 - 200mg/dL | 9 (22.5) | 31 (77.5) |  | 8 (44.4) | 10 (55.6) |  |
| ≥ 200mg/dL | 16 (22.9) | 54 (77.1) |  | 24 (29.6) | 57 (70.4) |  |
| **HbA1c, (n=114)** |  |  |  |  |  |  |
| < 7% | 3 (20.0) | 12 (80.0) | 0.988 <sup>a</sup> | 4 (23.5) | 13 (76.5) | 0.591 <sup>a</sup> |
| 7-9% | 4 (22.2) | 14 (77.8) |  | 15 (37.5) | 25 (62.5) |  |
| ≥ 9% | 17 (21.0) | 64 (79.0) |  | 24 (34.29) | 46 (65.7) |  |

Data are expressed as n (%) unless otherwise indicated. p-values were calculated using Pearson's chi-square test (<sup>a</sup>) or Fisher's exact test (<sup>b</sup>), as appropriate. "Other districts" include Surco, Barranco, Lurín, Pachacámac, San Bartolo, Pucusana, Punta Negra, Punta Hermosa, and Santa María del Mar. MDR-TB: multidrug-resistant tuberculosis (resistance to at least isoniazid and rifampicin). Any drug resistance includes MDR-TB, polyresistance, and monoresistance combined.

In the multivariate analysis, the prevalence of MDR-TB increased 1.9-fold in the youngest group aged 40 to 49 years and 3.1-fold in the oldest group aged 70 to 85 years (PR = 2.99; 95% CI = 1.00-8.97, p = 0.049) and (PR = 4.13; 95% CI = 1.16-14.7, p = 0.028), respectively. Compared to the central age group 50–59 years, which had the lowest prevalence and was used as the reference, individuals aged 40–49 and 70–85 showed significantly higher odds of MDR-TB, with (PR = 2.99;95% CI = 1.00–8.97, p=0.049) and (PR= 4.13; 95% CI= 1.16–14.7, p=0.028), respectively. These results were adjusted for age and body mass index (Table 4).

**Table 4.**
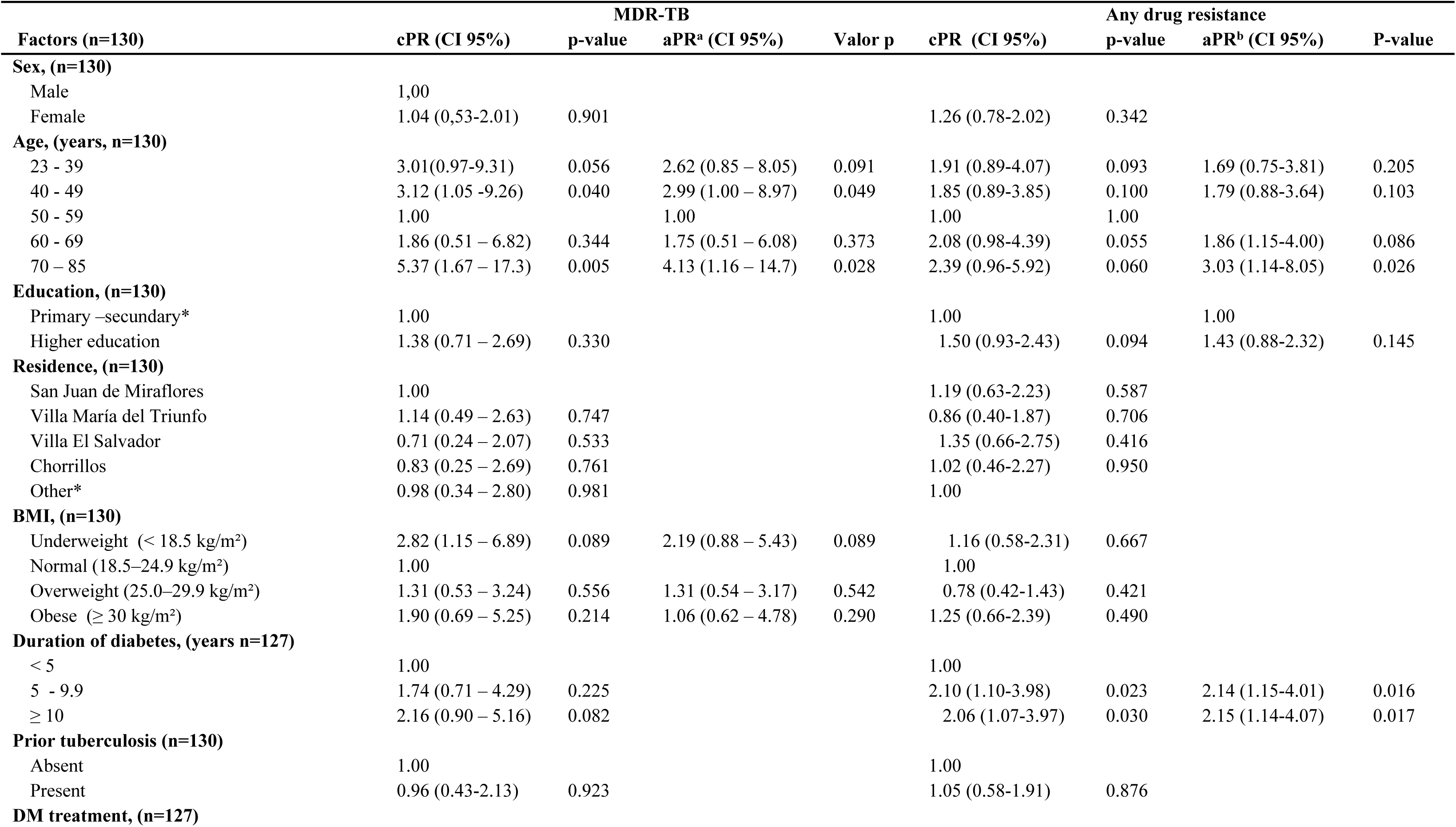
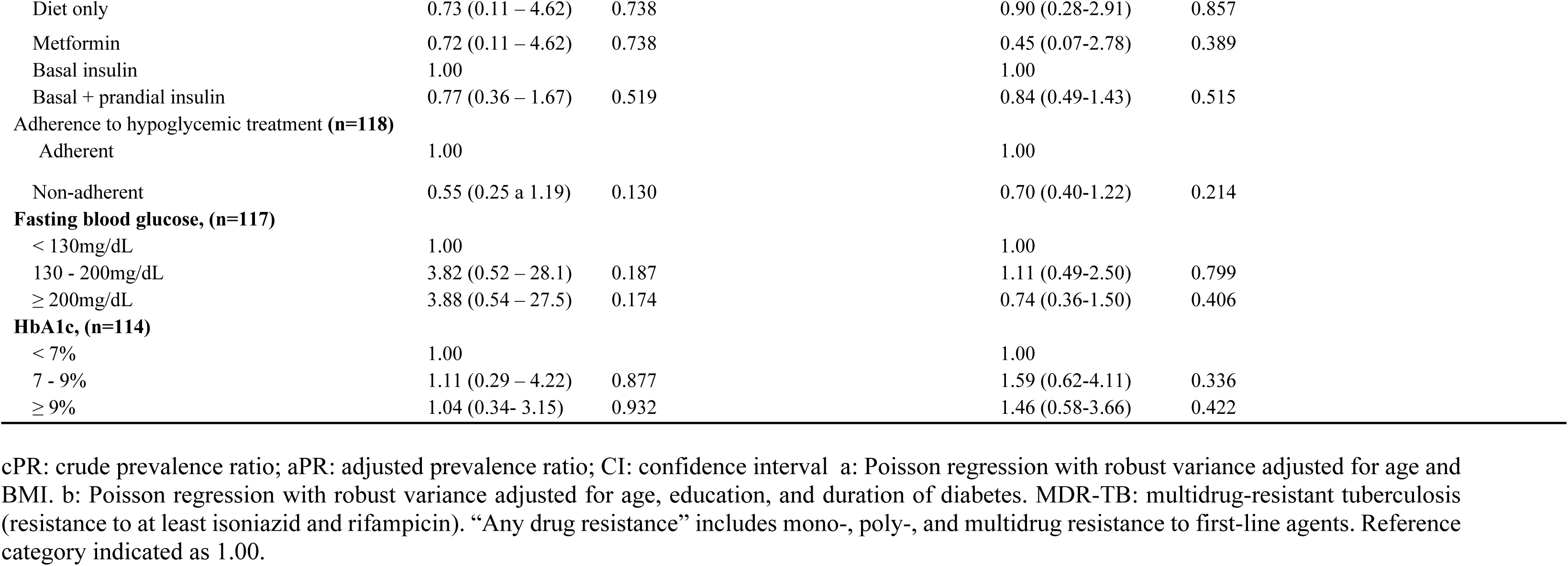
Multivariable Analysis of Factors Associated with Multidrug-Resistant Tuberculosis (MDR-TB) and Any Resistance to First-Line Drugs in Patients with Type 2 Diabetes Mellitus and Pulmonary Tuberculosis.

### Factors Associated with Bacterial Resistance to First-Line Drugs

In the bivariate analysis, DM duration of 5 years showed a high prevalence of resistance to some first-line drugs (p=0.031). There were no differences in the other variables (Table 3).

In the multivariate analysis, the groups with diabetes durations of 5-9 years and more than 10 years showed a 1.1-fold increase in resistance to some first-line drugs (PR = 2.14; 95% CI = 1.15-4.01, p=0.016) and (PR = 2.15; 95% CI= 1.14-4.07, p=0.017), compared to those with diabetes durations of less than 5 years. These results were adjusted for age, diabetes duration, and level of education. Similarly, resistance to certain first-line anti-tuberculosis drugs was nearly twice as prevalent among individuals aged 70–89 age group than in the 50–59 age group (PR 3.03; 95% CI= 1.14–8.05; p=0.026) **(Table 4).**

## Discussion

### Principal findings

This study documents a high frequency of bacterial resistance to first-line anti-tuberculosis drugs among patients with tuberculosis and type 2 diabetes mellitus (TB-DM) co-infection. A total of 34.6% of the cases exhibited resistance to at least one pharmaceutical agent, while 21.5% were multidrug-resistant tuberculosis (MDR). These proportions are much higher than those reported for the general tuberculosis population in Peru. Furthermore, significant associations were identified with age and duration of diabetes. Patients aged 40–49 and ≥70 years, as well as those with a duration of DM exceeding 5 years, exhibited an elevated risk of developing resistance. By contrast, there was no significant association between resistance and variables such as sex, glycemic control, history of previous tuberculosis, body mass index (BMI) and adherence to hypoglycemic treatment.

### Comparison with other studies

Various studies have documented disparate rates of anti-tuberculosis drug resistance among patients with TB-DM. In a Mexican cohort of patients with type 2 diabetes, 14% had MDR and 19% had resistance to at least one first-line drug (21). The prevalence of MDR in Indonesia was 17% (22). In china, the prevalence of MDR of 6.6% and resistance to at least one drug in 23% of cases has been reported. (23). Among patients with TB-DM, 15.6% had MDR and 9.1% had some resistance, as documented in Peru. (24). However, the populations, levels of care, and methodologies included in those studies could explain the differences with our research.

Previous studies have reported that the duration of DM is a predictor of MDR, as are a history of TB and poor glycemic control (25,26). In our study, duration of diabetes was significantly associated with resistance. However, no significant associations were found with a history of TB or HbA1c. This phenomenon can be explained by the high proportion of patients with previous TB (>80%) and poor glycemic control (87% with HbA1c ≥7%), which reduced the variability necessary to detect significant differences.

On the other hand, previous studies have pointed to a higher risk of resistance in young adults (aged 45-64), probably due to greater community exposure and lower therapeutic adherence (25). In contrast, our study identified a heightened risk among older adults (≥ 70 years old). This elevated risk may be attributable to a combination of factors, including a greater prevalence of comorbidities, immunological alterations associated with aging, and a history of exposure to TB or suboptimal treatment regimens.

### Immunological and Pharmacological mechanisms

The high prevalence of multidrug-resistant tuberculosis (MDR-TB) can be attributed to multiple clinical and biological factors. Firstly, the analysis revealed that most patients had a history of previous TB treatment, indicating a high proportion of retreatment cases. This finding suggests an increased likelihood of developing resistant strains (25).

From an immunological perspective, type 2 diabetes mellitus(T2DM) induces a state of chronic immunosuppression that is mediated by sustained hyperglycemia. This phenomenon affects multiple components of the immune system, including the dysfunction of neutophils, macrophages, and T lymphocytes, as well as alterations in the production of reactive oxygen species (ROS) (27). Furthermore, non-enzymatic glycation of proteins — including immunoglobins — can compromise the immune response to Mycobacterium tuberculosis, leading to an increased bacterial load, greater radiological extent, and a lower bacteriological conversion rate (28).

Pharmacokinetic studies suggest that patients with DM may exhibit reduced plasma concentracions of rifampicin and isoniazid. This reduction has been associated with factors such as diabetic gastroparesis, hepatic metabolic alterations and enhanced renal clearance (29). Moreover, interactions between medications such as that observed between rifampicin and metformin have been associated with impaired glycemic control, thereby inducing a cycle of hyperglycemia, immunosuppression and therapeutic ineffectiveness (30).

The association between advanced age and resistance may reflect age-related immune deterioration (immunosenescence), a higher burden of comorbidities, anda history of previous anti-tuberculosis treatment(25). Likewise, the prolonged nature of diabetes has been associated with cumulative immune damage, inadequate glycemic control, and impaired drug absorption or metabolism (31).

Despite the absence of a statistically significant association between elevated HbA1c and resistance, it is notable that 71% of patients with TB-DM exhibited HbA1c levels ≥9%, suggesting a potential role of hyperglycemia in the development of drug resistance. Previous studies have reported significant associations between HbA1c ≥9% and MDR-TB (25,26).

Although the association was not statistically significant, the U-shaped pattern observed between BMI and resistance suggests that both malnutrition and obesity may act as risk factors. Malnutrition may contribute by compromising immune function, white obesity may promote resistance trough chronic inflammation and insuline resistance.

### Adherence to hypoglycemic treatment

Treatment regimens for TB-DM are frequently rigid, which can result in a cycle of noncompliance that is further exacerbated by socioeconomic factors and associated stigma (32). In addition, patients with DM are more likely to experience adverse drug reactions, which may increase treatment rejection (33).

Poor treatment adherence in patients with TB-DM is associated with higher mortality and increased complications (34). Insulin is a key treatment in TB-DM due to its safety profile and glycemic efficacy, but limited access may reduce adherence in low-resource settings. Therefore, it should be complemented with interventions targeting diet and physical activity to minimize pharmacological dependence and improve TB outcomes (35).

Comprehensive care models that monitor TB and diabetes can improve comorbidity managment, but remain difficult to implement in fragile, centralized health systems (36).

### Implications for Public Health

The rising burden of TB-diabetes comorbidity threatens public health, particularly in low- and middle-income countries. DM increases the risk of developing active TB by two to three times and is linked to worse clinical outcomes, including higher rates of treatment failure, relapse and drug resistance (37,38).

Our findings highlight the importance of integrating tuberculosis and noncommunicable disease control programs. This includes bidirectional screening, glycemic control during TB treatment, and optimizing regimens for high-risk patients. The dual burden of TB-DM generates substantially higher healthcare costs. These challenges contribute to greater risk of treatment abandonment and socioeconomic inequity in affected populations (39).

#### Strengths and limitations

This study has several limitations. Its retrospective cross-sectional design is primarily descriptive and aimed at estimating prevalences; therefore, any associations evaluated should be interpreted as exploratory. Additionally, missing data on key variables such as HbA1c, treatment adherence, and glycemic control limited further analyses, including assessment of plasma drug concentrations and pharmacodynamic responses.

Nevertheless, the study has several strengths. It included patients treated at a specialized referral center for drug-resistant tuberculosis and comorbidities, enhancing the clinical relevance of the findings. Laboratory diagnoses were based on bacterial sensitivity tests validated by Peruvian National Institute of Health, ensuring methodological reliability. Finally, adjusted multivariate analysis clarified resistance patterns in TB-DM under high-burden settings. Moreover, including all eligible cases through a census approach minimized sampling error and enhanced the internal validity of the study findings.

### Research recommendations

Prospective studies are needed to assess whether intensive glycemic control improves TB treatmet outcomes. Pharmacokinetic evaluations should clarify if dose adjustments or individualizad rifampicin and isoniazid regimens are warranted in patients with diabetes. Further research should clarify immune responses in TB-DM comorbidit — macrophage function, T-cell activity and cytokine profiles — and advance genomic tools to distinguish acquired from transmitted MDR-TB. A crucial aspect is the evaluation of integrated public health strategies, like combined TB-DM clinics and patient programs. These interventions aim to strengthen therapeutic adherence and support self-care.

## Conclusions

TB-DM appears to increase the risk of resistance to first-line anti-tuberculosis drugs, including MDR-TB. Older age and longer duration of diabetes may contribute to this association. The mechanisms involved include immunosuppression mediated by hyperglycemia, pharmacokinetic alterations, and cumulative exposure to previous treatments.

These findings support the integration of TB and DM programs, with emphasis on surveillance, treatment optimization, and targeted follow-up in high-risk groups. A syndemic approach to TB-DM is essential to reduce the burden of disease, improve patient prognosis, and contain the spread of resistant tuberculosis.

## Data Availability

All relevant data are within the manuscript and its Supporting Information files.

## References

1. Obermeyer Z, Abbott-Klafter J, Murray CJL. Has the DOTS strategy improved case finding or treatment success? An empirical assessment. PLoS One. 2008;3(3):e1721. doi: 10.1371/journal.pone.0001721.

2. Kumar V, Khatib MN, Verma A, Lakhanpal S, Ballal S, Kumar S, et al. Tuberculosis in South Asia: A regional analysis of burden, progress, and future projections using the global burden of disease (1990–2021). J Clin Tuberc Other Mycobact Dis. 2024;37:100480. doi: 10.1016/j.jctube.2024.100480.

3. Lv H, Wang L, Zhang X, Dang C, Liu F, Zhang X, et al. Further analysis of tuberculosis in eight high-burden countries based on the Global Burden of Disease Study 2021 data. Infect Dis Poverty. 2024;13:70. doi: 10.1186/s40249-024-01247-8.

4. Chen Z, Wang T, Du J, Sun L, Wang G, Ni R, et al. Decoding the WHO Global Tuberculosis Report 2024: A critical analysis of global and Chinese key data. Zoonoses. 2025;5:1. doi: 10.15212/ZOONOSES-2024-0061.

5. Yang H, Ruan X, Li W, Xiong J, Zheng Y. Global, regional, and national burden of tuberculosis and attributable risk factors for 204 countries and territories, 1990–2021: a systematic analysis for the Global Burden of Diseases 2021 study. BMC Public Health. 2024;24:3111. doi: 10.1186/s12889-024-20664-w.

6. Villalva-Serra K, Barreto-Duarte B, Rodrigues MM, Queiroz ATL, Martinez L, Croda J, et al. Impact of strategic public health interventions to reduce tuberculosis incidence in Brazil: a Bayesian structural time-series scenario analysis. Lancet Reg Health Am. 2025;41:100963. Epub 2024 Dec 11. doi: 10.1016/j.lana.2024.100963.

7. de Paiva JPS, Magalhães MAFM, Leal TC, da Silva LF, da Silva LG, do Carmo RF, et al. Time trend, social vulnerability, and identification of risk areas for tuberculosis in Brazil: An ecological study. PLoS One. 2022;17(1):e0247894. doi: 10.1371/journal.pone.0247894.

8. Suárez PG, Watt CJ, Alarcón E, Portocarrero J, Zavala D, Canales R, et al. The dynamics of tuberculosis in response to 10 years of intensive control effort in Peru. J Infect Dis. 2001;184(4):473–478. Epub 2001 Jul 18. doi: 10.1086/322777.

9. Ministerio de Salud (MINSA), Dirección de Redes Integradas de Salud Lima Este. Análisis de la situación epidemiológica de la tuberculosis en DIRIS Lima Este 2024 [Internet]. Lima: MINSA; 2024 [cited 2025 Sep 22]. Available from: https://cdn.www.gob.pe/uploads/document/file/7989640/6715867-analisis-de-la-situacion-epidemiologica-de-la-tuberculosis-de-la-direccion-de-redes-integradas-de-salud-lima-este-2024.pdf?v=1745854892

10. Wingfield T, Boccia D, Tovar M, Gavino A, Zevallos K, Montoya R, et al. Defining catastrophic costs and comparing their importance for adverse tuberculosis outcome with multi-drug resistance: A prospective cohort study, Peru. PLoS Med. 2014;11(7):e1001675. doi: 10.1371/journal.pmed.1001675.

11. Walter KS, Martinez L, Arakaki-Sanchez D, Sequera VG, Estigarribia Sanabria G, Cohen T, et al. The escalating tuberculosis crisis in central and South American prisons. Lancet. 2021;397(10284):1591–1596. Epub 2021 Apr 8. doi: 10.1016/S0140-6736(20)32578-2.

12. Edessa D, Sisay M, Dessie Y. Unfavorable outcomes to second-line tuberculosis therapy among HIV-infected versus HIV-uninfected patients in sub-Saharan Africa: A systematic review and meta-analysis. PLoS One. 2020;15(8):e0237534. doi: 10.1371/journal.pone.0237534.

13. Montalvo-Otivo R, Ramírez-Breña M, Bruno-Huamán A, Damián-Mucha M, Vilchez-Bravo S, Quisurco-Cárdenas M. Distribución geográfica y factores de riesgo de tuberculosis multidrogorresistente en el centro de Perú. Rev Fac Med. 2020;68(2):245–250. doi: 10.15446/revfacmed.v68n2.71715.

14. Villegas L, Otero L, Sterling TR, Huaman MA, Van der Stuyft P, Gotuzzo E, et al. Prevalence, risk factors, and treatment outcomes of isoniazid- and rifampicin-mono-resistant pulmonary tuberculosis in Lima, Peru. PLoS One. 2016;11(4):e0152933. doi: 10.1371/journal.pone.0152933.

15. Jeon CY, Murray MB. Diabetes mellitus increases the risk of active tuberculosis: A systematic review of 13 observational studies. PLoS Med. 2008;5(7):e152. doi: 10.1371/journal.pmed.0050152.

16. Nathella PK, Babu S. Influence of diabetes mellitus on immunity to human tuberculosis. Immunology. 2017;152(1):13–24. Epub 2017 Jun 29. doi: 10.1111/imm.12762.

17. Vera-Ponce VJ, Zuzunaga-Montoya FE, Vásquez-Romero LEM, Loayza-Castro JA, Vigil-Ventura E, Ramos W. Prevalence of diabetes and prediabetes in Peru: a systematic review and meta-analysis. Diabetol Metab Syndr. 2025;17(1):260. doi: 10.1186/s13098-025-01844-z.

18. Wu Q, Liu Y, Ma Y-B, Liu K, Chen S-H. Incidence and prevalence of pulmonary tuberculosis among patients with type 2 diabetes mellitus: A systematic review and meta-analysis. Ann Med.2022;54(1):1657–1666.doi: 10.1080/07853890.2022.2085318.

19. Magee MJ, Bloss E, Shin SS, Contreras C, Arbanil Huaman H, Calderon Ticona J, et al. Clinical characteristics, drug resistance, and treatment outcomes among tuberculosis patients with diabetes in Peru. Int J Infect Dis. 2013;17(6):e404–e412. Epub 2013 Feb 22. doi: 10.1016/j.ijid.2012.12.029.

20. Servicio Galego de Saúde (SERGAS), Consellería de Sanidade, Xunta de Galicia. Epidat 4.2 [Internet]. Santiago de Compostela (ES): SERGAS; 2016– [cited 2025 Sep 22]. Available from: https://www.sergas.es/saude-publica/epidat-4-2?idioma=es

21. Perez-Navarro LM, Restrepo BI, Fuentes-Dominguez FJ, Duggirala R, Morales-Romero J, López-Alvarenga JC, et al. The effect size of type 2 diabetes mellitus on tuberculosis drug resistance and adverse treatment outcomes. Tuberculosis (Edinb). 2017;103:83–91. Epub 2017 Jan 24. doi: 10.1016/j.tube.2017.01.006.

22. Saktiawati AMI, Subronto YW. Influence of diabetes mellitus on the development of multi drug resistant-tuberculosis in Yogyakarta. Acta Med Indones. 2018;50(1):11–17. Epub 2018 Jan.

23. Wu Q, Wang M, Zhang Y, Wang W, Ye T-F, Liu K, Chen S-H. Epidemiological characteristics and their influencing factors among pulmonary tuberculosis patients with and without diabetes mellitus: A survey study from drug resistance surveillance in East China. Front Public Health. 2022;9:777000. Epub 2022 Jan 24. doi: 10.3389/fpubh.2021.777000.

24. Magee MJ, Bloss E, Shin SS, Contreras C, Arbanil Huaman H, Calderon Ticona J, et al. Clinical characteristics, drug resistance, and treatment outcomes among tuberculosis patients with diabetes in Peru. Int J Infect Dis. 2013;17(6):e404–e412. Epub 2013 Feb 22. doi: 10.1016/j.ijid.2012.12.029.

25. Li S, Liang Y, Hu X. Risk factors for multidrug resistance in tuberculosis patients with diabetes mellitus. BMC Infect Dis. 2022;22(1):835. doi: 10.1186/s12879-022-07831-3.

26. Lyu M, Wang D, Zhao J, Yang Z, Chong W, Zhao Z, et al. A novel risk factor for predicting anti-tuberculosis drug resistance in patients with tuberculosis complicated with type 2 diabetes mellitus. Int J Infect Dis. 2020;97:69–77. doi: 10.1016/j.ijid.2020.05.080.

27. Aravindhan V, Yuvaraj S. Immune-endocrine network in diabetes–tuberculosis nexus: does latent tuberculosis infection confer protection against meta-inflammation and insulin resistance? Front Endocrinol (Lausanne). 2024;15:1303338. doi: 10.3389/fendo.2024.1303338.

28. Daryabor G, Atashzar MR, Kabelitz D, Meri S, Kalantar K. The Effects of Type 2 Diabetes Mellitus on Organ Metabolism and the Immune System. Front Immunol. 2020;11:1582. doi: 10.3389/fimmu.2020.01582.

29. Zhu Y, Davies Forsman L, Chen C, Zhang H, Shao G, Wang S, et al. Drug exposure and treatment outcomes in patients with multidrug-resistant tuberculosis and diabetes mellitus: a multicenter prospective cohort study from China. Clin Infect Dis. 2024;79(2):524–533. doi: 10.1093/cid/ciae329.

30. Rissaadah S, Nursiswati N, Pahria T. Glycemic profile and clinical treatment in patients with diabetes mellitus–tuberculosis: An update scoping review. J Multidiscip Healthc. 2025;18:747–758. doi: 10.2147/JMDH.S510247.

31. Xu G, Hu X, Lian Y, Li X. Diabetes mellitus affects the treatment outcomes of drug-resistant tuberculosis: a systematic review and meta-analysis. BMC Infect Dis. 2023;23(1):813. doi: 10.1186/s12879-023-08765-0.

32. Murwanashyaka JD, Ndagijimana A, Biracyaza E, Sunday FX, Umugwaneza M. Non-adherence to medication and associated factors among type 2 diabetes patients at Clinique Medicale Fraternite, Rwanda: a cross-sectional study. BMC Endocr Disord. 2022;22(1):219. doi: 10.1186/s12902-022-01133-0.

33. Mistry J, Sinha A, Divakar B, Gavli N, Vadgama P. An observational study on the outcome of antitubercular and antidiabetic therapy in patients of tuberculosis with diabetes mellitus as comorbidity. Asian J Pharm Clin Res. 2024;17(7):113–116. doi: 10.22159/ajpcr.2024v17i7.50922.

34. Degner NR, Wang J-Y, Golub JE, Karakousis PC. Metformin use reverses the increased mortality associated with diabetes mellitus during tuberculosis treatment. Clin Infect Dis. 2018;66(2):198–205. doi: 10.1093/cid/cix819.

35. Godwin IU, Atulomah N. Impact of lifestyle change intervention on tuberculosis treatment outcome in tuberculosis patients with diabetes mellitus comorbidity in South West Nigeria. Texila International Journal of Public Health. 2023;11(2):Art023. doi: 10.21522/TIJPH.2013.11.02.Art023.

36. Udaykumar P, Kumar S, Chandralekha N, Reddy RH, Badarudeen MN, Nagaraja SB. Daily monitoring of diabetic treatment amongst TB-DM patients under NTEP: Does it improve the treatment outcomes? Clin Epidemiol Glob Health. 2022;17:101118. doi: 10.1016/j.cegh.2022.101118.

37. Jeon CY, Murray MB. Diabetes mellitus increases the risk of active tuberculosis: A systematic review of 13 observational studies. PLoS Med. 2008;5(7):e152. doi: 10.1371/journal.pmed.0050152.

38. Rehman AU, Khattak M, Mushtaq U, Latif M, Ahmad I, Rasool MF, et al. The impact of diabetes mellitus on the emergence of multi-drug resistant tuberculosis and treatment failure in TB-diabetes comorbid patients: a systematic review and meta-analysis. Front Public Health. 2023;11:1244450. doi: 10.3389/fpubh.2023.1244450.

39. Yamanaka T, Castro MC, Ferrer JP, Solon JA, Cox SE, Laurence YV, et al. Costs incurred by people with co-morbid tuberculosis and diabetes and their households in the Philippines. PLoS One. 2024;19(1):e0297342. doi: 10.1371/journal.pone.0297342.

